# Relationship between dietary choline intake and diabetes mellitus in the National Health and Nutrition Examination Survey 2007-2010

**DOI:** 10.1101/2020.08.09.20171306

**Authors:** Long Zhou, Xiang Li, Shuhong Li, Xiaoxiao Wen, Yaguang Peng, Liancheng Zhao

## Abstract

**Background:** Previous studies have shown that elevated trimethylamine N-oxide (TMAO) was associated with a higher risk of diabetes mellitus (DM). Little is known about the relationship between dietary intake of choline, which is a major dietary precursor for gut microbiome-derived TMAO, and DM in the general population.

**Objective:** The present study aims to explore the relationship between dietary choline intakes and DM in the United States (US) adult population.

**Design:** Cross-sectional data were derived from the National Health and Nutrition Examination Survey (NHANES) 2007-2010 of 8621 individuals aged 20 years or older. Multivariable logistic regression models were used to determine odds ratios (ORs) and 95% confidence intervals (CIs) for DM of each quartile category of energy-adjusted choline intakes. The restricted cubic spline model was used for the dose-response analysis. The receiver operating characteristic (ROC) curve was used to determine the optimal cut-off value of choline intake for predicting DM.

**Results:** A linear dose-response relationship between dietary choline intakes and the odds of DM was found after adjustment for multiple potential confounding factors, p for linear =0.0002. With the lowest quartile category of choline as the reference, the multivariable-adjusted ORs and 95% CIs of the second, third, and highest quartile categories were 1.22 (0.98, 1.52), 1.26 (1.01, 1.56), and 1.42 (1.15, 1.77), respectively, p for trend =0.0024. Per 100 mg/d increase in energy-adjusted choline resulted in 15% (95% CI: 7%, 22%) higher odds of DM. The ROC analysis identified an energy-adjusted choline of 331.7 mg/d as the optimal cut-off value for predicting DM, with 52.5% sensitivity and 60.7% specificity.

**Conclusion:** This study supports a positive and linear relationship between dietary choline intake and DM in the US adult population. Further studies are warranted to replicate our findings in other populations and elucidate the potential mechanisms.

## INTRUODUCTION

Diabetes mellitus (DM), a heterogeneous mix of health conditions characterized by glucose dysregulation, is a major risk factor for cardiovascular disease (CVD), including coronary heart disease and stroke (1). DM confers about a two-fold excess risk for a wide range of CVD, independently from other conventional risk factors (2). According to the International Diabetes Federation (IDF) Atlas, the global DM prevalence is estimated to be 9.3% (463 million people) in 2019, and is predicted to rise to 10.2% (578 million) by 2030 and 10.9% (700 million) by 2045 (3). Diet is an important modifiable factor involved in the development of cardiometabolic diseases such as hypertension (4) and DM (5).

Choline is a major dietary precursor for gut microbiome-derived trimethylamine, which is converted in the liver to trimethylamine N-oxide (TMAO) by hepatic enzyme flavin-containing monooxygenases (6). Previous evidence has shown that elevated TMAO was associated with a higher risk of CVD (7,8) and DM (9,10). However, little is known about the relationship between dietary choline intake and DM in the general population and results from previous studies are controversial (11,12). In this context, we aimed to explore the relationship between dietary choline intake and DM in the United States (US) adult population.

## METHODS

### Study population

Data were derived from the National Health and Nutrition Examination Survey (NHANES) 2007-2010. A detailed description of the NHANES study design and methods are available elsewhere (13). Briefly, NHANES is a program of ongoing cross-sectional surveys that are conducted by the National Center for Health Statistics (NCHS), which is a part of the Centers for Disease Control and Prevention (CDC). A major objective of NHANES is to estimate the number and percentage of people with selected diseases and risk factors in the US population. In total, 20686 individuals completed the interviews in the NHANES 2007-2010 cohort. In this study, we included nonpregnant individuals aged ≥20 y who completed two 24-h recalls with reliable energy intakes (n=9371). The reliable energy intake was defined as total energy intake from any 24-h recall ranges from 500 to 5000 kcal/day for women and from 500 to 8000 kcal/day for men (4). We further excluded 750 subjects for missing data on Hemoglobin A1c (HbA1c) or covariates such as waist circumference (WC), body mass index (BMI), and C-reactive protein (CRP). This resulted in 8621 participants included in the final analysis. A detailed flow chart depicting participant selection is shown in **Figure 1**. The NCHS institutional review board approved the study protocol, and all participants provided written informed consent. All data used in this manuscript are de-identified and freely available to the public through: https://www.n.cdc.gov/nchs/nhanes/default.aspx.

**Figure 1.**
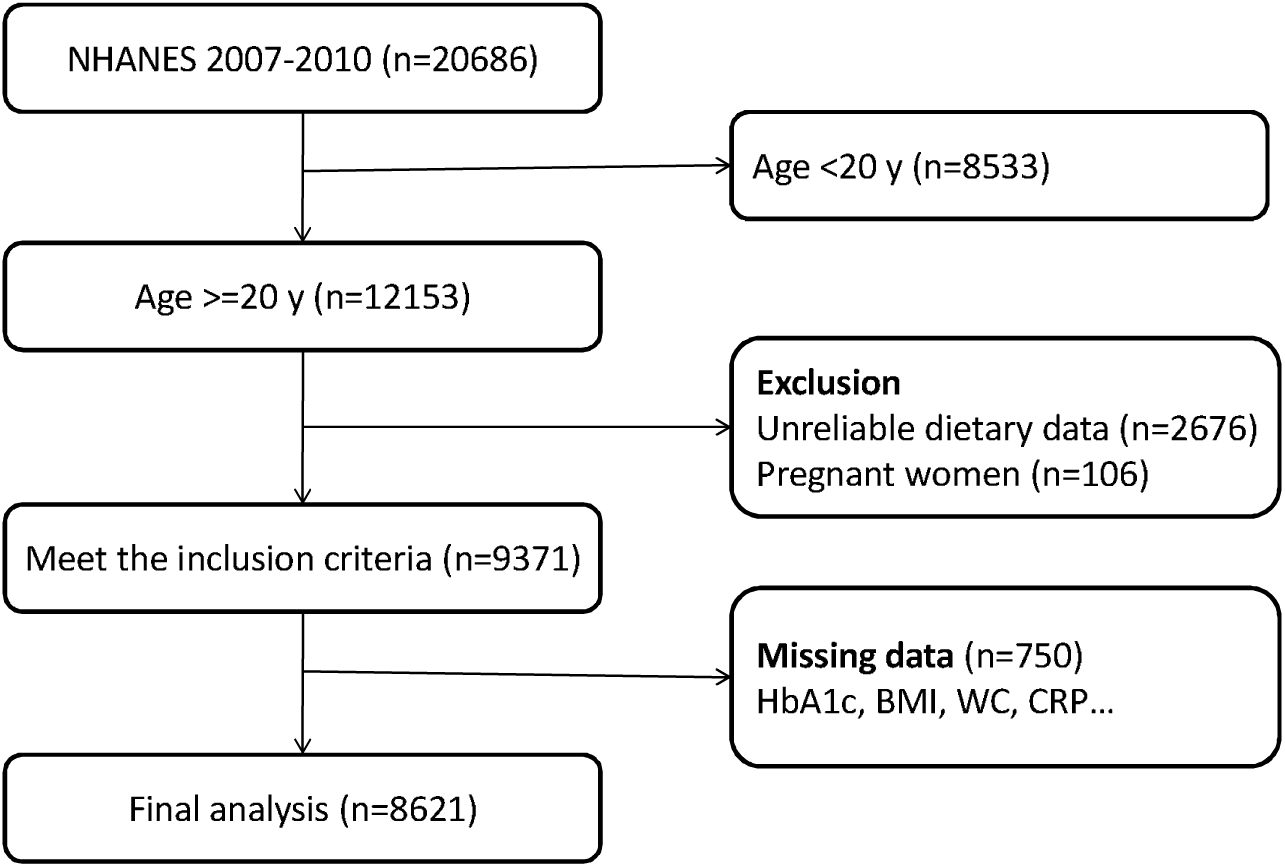
Flowchart of participant selection

### Data Collection

Data were collected during interviews in the participants’ homes and through medical examination and subsequent laboratory assessments in the Mobile Examination Centre. The NHANES interviews were completed by trained interviewers in participants’ homes by using a Computer-Assisted Personal Interview (CAPI) system. The questionnaire contains a number of questions on demographics, health conditions, and lifestyles such as smoking, drinking, and passive sedentary behavior.

The NHANES 2007-2010 ethnicity variable included Mexican American, other Hispanic, non-Hispanic White, non-Hispanic Black, and other races. To facilitate the analysis, we merged Mexican American and other Hispanic into one group (Hispanic). The 2007 to 2010 Department of Health and Human Services’ (HHS) poverty guidelines were used to calculate family monthly poverty level index by dividing family income by the poverty guidelines, specific to family size, as well as the appropriate year and state. The index was then grouped into three categories (<=1.30, 1.31~1.85, and >1.85) and we used the family monthly poverty level category as the house income variable in the present analysis.

With respect to lifestyle variables, smoking status was categorized into current, former, and never smoker according to participants’ answers to the questionnaire. Participants drinking alcohol at least 12 times in the previous year were defined as drinkers. Total time spent sitting in a typical day not including time spent sleeping was used as a passive sedentary behavior indicator in this analysis.

Body weight (kg) was measured using a digital weight scale and standing height (cm) was measured using a stadiometer. BMI (kg/m^2^) was calculated as weight in kilograms divided by height in metres squared, and then rounded to one decimal place.

The dietary interview component, called What We Eat in America (WWEIA), is conducted as a partnership between the US Department of Agriculture (USDA) and the US Department of Health and Human Services (DHHS). Under this partnership, DHHS’ National Center for Health Statistics (NCHS) is responsible for the survey sample design and all aspects of data collection and USDA’s Food Surveys Research Group (FSRG) is responsible for the dietary data collection methodology, maintenance of the databases used to code and process the data, and data review and processing. NHANES participants are eligible for two 24-hour dietary recall interviews. The first dietary recall interview was collected face-to-face in the Mobile Examination Centre (MEC), and the second interview was collected by telephone 3 to 10 days later. Dietary intake was estimated as a mean of the two dietary recalls. Energy-adjusted dietary choline intake (mg per 2000 kcal) was used as the main exposure variable. Total energy, saturated fatty acids (SFA), polyunsaturated fatty acids (PFA), fiber, as well as nutrients involved in the choline metabolism such as dietary folate, vitamin B-6, and vitamin B-12 intakes were included as potential confounding factors in the analysis.

### Laboratory tests and clinical definition

Blood specimens were processed, stored and shipped to Fairview Medical Center Laboratory at the University of Minnesota, Minneapolis Minnesota for analysis. The analyzer diluted the whole blood specimen with a hemolysis solution, and then injected a small volume of the treated specimen onto the HPLC analytical column. Separation was achieved by utilizing differences in ionic interactions between the cation exchange group on the column resin surface and the hemoglobin components. The hemoglobin fractions (A1c, A1b, F, LA1c, SA1c, A0 and H-Var) were subsequently removed from the column material by step-wise elution using elution buffers each with a different salt concentration. The separated hemoglobin components passed through the photometer flow cell where the analyzer measures changes in absorbance at 415 nm. The analyzer integrated and reduced the raw data, and then calculated the relative percentages of each hemoglobin fraction. We defined diabetes on the basis of HbA1c ≥6.5% and/or current treatment with a hypoglycemic agent or insulin (14). CRP was measured by latex-enhanced nephelometry on a Behring Nephelometer.

### Statistical analysis

Data were presented as mean ± SD for continuous variables and as percentages for categorical variables. We tested differences in characteristics between groups with student’s t-tests for continuous variables, and with χ2 tests for categorical variables. Dietary choline intake was analyzed both as a continuous and categorical variable (quartiles). When analyzed continuously, choline intake was rescaled to units of 100 mg/d. The odds ratios (ORs) and 95% confidence intervals (CIs) for DM of each choline category and each unit increase in choline were estimated using multivariate non-conditional logistic regression models. To test for trends across quartile of choline intake, we modelled the median values of each quartile as a continuous variable in all models. Potential covariates such as age, sex, ethnicity, education levels, family monthly poverty levels, passive sedentary hours, smoking status, drinking status, BMI, CRP, total energy, SFA, PFA, fiber dietary folate, vitamin B-6, and vitamin B-12, were included in the full-adjusted models. The restricted cubic spline model was used for the dose–response analysis. We also used receiver operating characteristic (ROC) curve to determine the optimal cut-off value of choline intake for predicting DM. We divided participants into two groups according to their choline intake below or above the optimal cut-off value and further calculated the OR of the higher choline intake group in comparison with the lower group. A two-tailed *P* < 0.05 was considered significant. All analyses were performed using SAS version 9.4 (SAS Institute, Cary, North Carolina) and R version 3.6.3 (R Foundation for Statistical Computing,Vienna, Austria).

## RESULTS

The characteristics of the study population are presented in **Table 1**. A total of 8621 participants (4199 men and 4422 women) with a mean ± SD age of 50.4 ± 17.5 years were included in this analysis. DM was diagnosed in 1093 (12.7%) participants. Compared with those without DM, participants with DM were older, and more likely to be of white race, to have higher levels of BMI, WC, and CRP, to have higher intakes of choline, SFA, PFA, fiber, folate, and vitamin B-12.

**Table 1.** Characteristics of the study population (n=8621)^1^

| Characteristics | All | Diabetes |  | P values |
| --- | --- | --- | --- | --- |
|  |  | No | Yes |  |
| n | 8621 | 7528 | 1093 |  |
| Age, y | 50.4±17.5 | 48.8±17.5 | 61.3±13.0 | <0.0001 |
| Sex, n(%) |  |  |  | 0.1612 |
| Men | 4199 (48.7) | 3645 (48.4) | 554 (50.7) |  |
| Women | 4422 (51.3) | 3883 (51.6) | 539 (49.3) |  |
| Ethnicity, n(%) |  |  |  | <0.0001 |
| Hispanic | 2414 (28.0) | 2062 (27.4) | 352 (32.2) |  |
| White | 4369 (50.7) | 3926 (52.2) | 443 (40.5) |  |
| Black | 1493 (17.3) | 1236 (16.4) | 257 (23.5) |  |
| Others | 345 (4.0) | 304 (4.0) | 41 (3.8) |  |
| Education categories, n(%) |  |  |  | <0.0001 |
| Less than 5th grade | 2356 (27.3) | 1932 (25.7) | 424 (38.8) |  |
| 5th to 8th grade | 2066 (24.0) | 1805 (24.0) | 261 (23.9) |  |
| More than or equal to 9th grade | 4199 (48.7) | 3791 (50.4) | 408 (37.3) |  |
| Family monthly poverty levels, n(%) |  |  |  | 0.0454 |
| ≤1.30 | 2779 (32.2) | 2403 (31.9) | 376 (34.4) |  |
| 1.31~1.85 | 1556 (18.1) | 1344 (17.9) | 212 (19.4) |  |
| >1.85 | 4286 (49.7) | 3781 (50.2) | 505 (46.2) |  |
| Drinking status, n(%) |  |  |  | <0.0001 |
| Yes | 5957 (69.1) | 5302 (70.4) | 655 (59.9) |  |
| No | 2664 (30.9) | 2226 (29.6) | 438 (40.1) |  |
| Smoking status, n(%) |  |  |  | <0.0001 |
| Current | 1795 (20.8) | 1608 (21.4) | 187 (17.1) |  |
| Former | 2223 (25.8) | 1846 (24.5) | 377 (34.5) |  |
| Never | 4603 (53.4) | 4074 (54.1) | 529 (48.4) |  |
| Body mass index, kg/m<sup>2</sup> | 29.1±6.5 | 28.5±6.3 | 32.9±7.0 | <0.0001 |
| Passive sedentary hours, h/d | 5.2±3.3 | 5.2±3.3 | 5.3±3.2 | 0.5322 |
| C-reactive protein, mg/dl | 0.4±0.8 | 0.4±0.8 | 0.6±0.8 | <0.0001 |
| Total energy intake, kcal/d | 2028.0±783.5 | 2060.9±790.9 | 1801.4±689.6 | <0.0001 |
| Energy-adjusted choline, mg/d | 326.2±109.9 | 321.5±107.4 | 358.6±121.4 | <0.0001 |
| Energy-adjusted SFA, g/d | 23.8±6.9 | 23.7±6.9 | 24.5±6.9 | 0.0005 |
| Energy-adjusted PFA, g/d | 16.2±5.6 | 16.1±5.5 | 16.8±5.5 | 0.0002 |
| Energy-adjusted fiber, g/d | 17.1±7.7 | 16.9±7.6 | 18.4±7.8 | <0.0001 |
| Energy-adjusted folate, dietary folate equivalents | 541.1±263.6 | 538.5±264.5 | 558.9±256.5 | 0.0170 |
| Energy-adjusted vitamin B-6, mg/d | 2.1±1.0 | 2.1±1.0 | 2.1±0.9 | 0.0869 |
| Energy-adjusted vitamin B-12, µg/d | 5.3±5.0 | 5.2±4.2 | 6.1±8.7 | <0.0001 |

<sup>1</sup> Values are presented as mean ± SD unless otherwise indicated.
Abbreviations: SFA, saturated fatty acids; PFA, polyunsaturated fatty acids

**Table 2** presents the prevalence of DM and the association between DM and energy-adjusted choline as both categorical and continuous variables separately. The prevalence of DM in the 4 quartile categories of dietary choline were 8.4%, 11.9%, 13.4%, and 17.1%, respectively, p for trend <0.0001. In the age- and sex-adjusted model, a higher level of choline was found to be positively associated with higher odds for DM, taking the lowest quartile of choline as the referent category. This association was not altered after further adjustment for several other covariates including ethnicity, education levels, family poverty levels, passive sedentary time, smoking status, drinking status, BMI, CRP, dietary energy, SFA, PFA, fiber, folate, vitamin B-6, and vitamin B-12 intakes. The multivariable-adjusted ORs and 95% CIs of the second, third, and highest quartile categories were 1.22 (0.98, 1.52), 1.26 (1.01, 1.56), and 1.42 (1.15, 1.77), respectively, p for trend =0.0024. Per unit (100 mg) increase in energy-adjusted choline resulted in 15% (95% CI: 7%, 22%) higher odds of DM when choline was analyzed as a continuous variable. Dose-response analysis revealed a significant linear relationship between choline and the odds of DM, p for linear =0.0002 (**Figure 2**).

**Figure 2.**
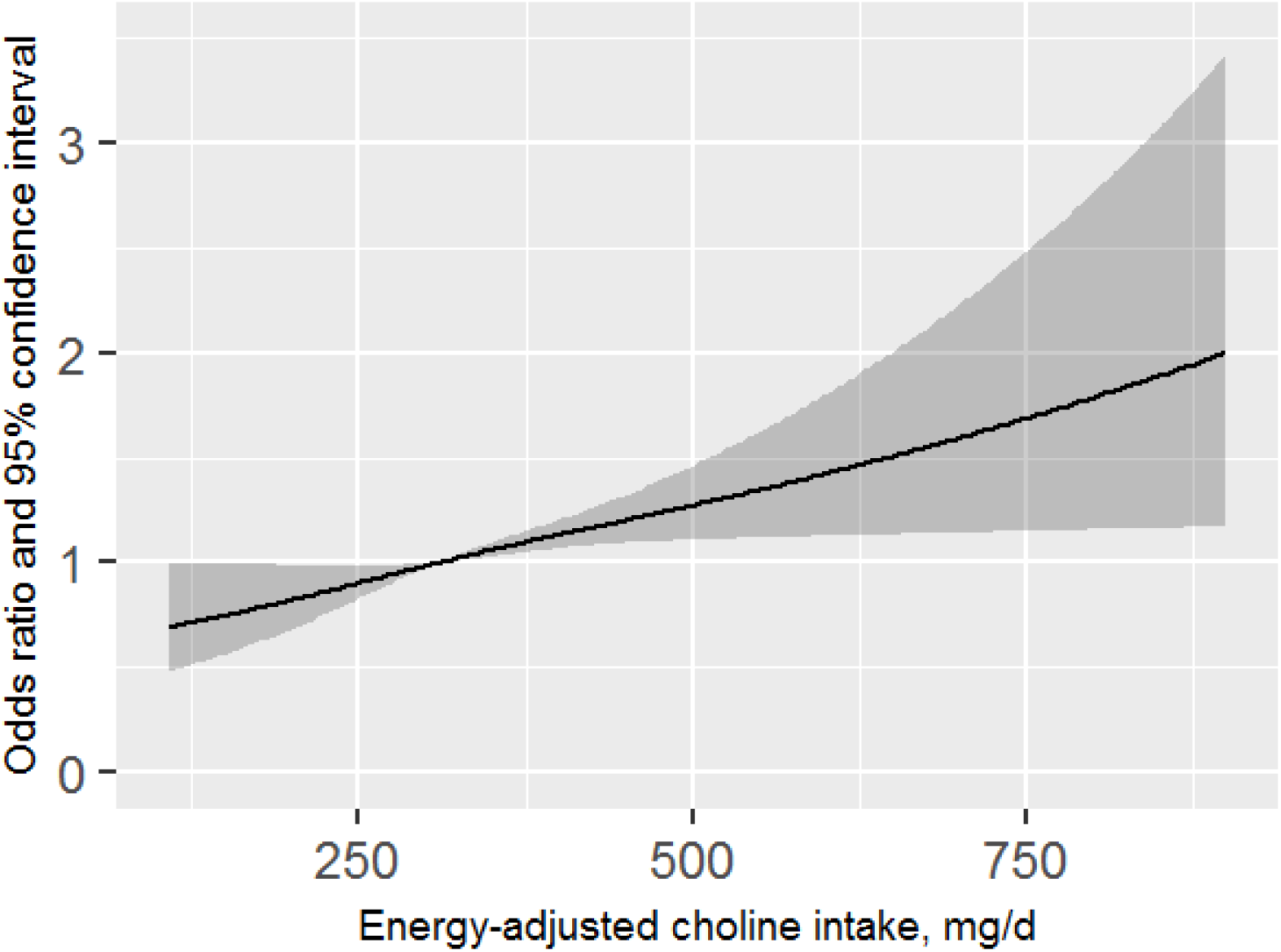
Dose-response relation between energy-adjusted choline intake and diabetes (P for linear =0.0002)

**Table 2.** Odds ratios and 95% confidence intervals for diabetes according to energy-adjusted choline intake levels

| Dietary choline | Diabetes/n | Prevalence (%) | Age- and sex-adjusted model | Full-adjusted model<sup>2</sup> |
| --- | --- | --- | --- | --- |
| Categorical<sup>1</sup> |  |  |  |  |
| Quartile 1 | 181/2158 | 8.4 | 1 (Ref) | 1 (Ref) |
| Quartile 2 | 256/2155 | 11.9 | 1.26 (1.03, 1.55) | 1.22 (0.98, 1.52) |
| Quartile 3 | 288/2152 | 13.4 | 1.30 (1.06, 1.59) | 1.26 (1.01, 1.56) |
| Quartile 4 | 368/2156 | 17.1 | 1.69 (1.39, 2.06) | 1.42 (1.15, 1.77) |
| P for trend |  | <0.0001 | <0.0001 | 0.0024 |
| Continuous |  |  |  |  |
| Per 100-mg/d increase in choline | 1093/8621 | 12.7 | 1.22 (1.16, 1.30) | 1.15 (1.07, 1.22) |

1 The quartile cut-off values of dietary choline are 249.6, 309.0, and 383.3 mg.
2 Full-adjusted model adjusted for age, sex, ethnicity, education levels, family income levels, smoking status, drinking status, passive sedentary hours, body mass index, C-reactive protein, dietary energy, saturated fatty acids, polyunsaturated fatty acids, fiber, folate, vitamin B-6, and vitamin B-12 intakes.

The ROC analysis identified an energy-adjusted choline of 331.7 mg/d as the optimal cut-off value for predicting DM, with 52.5% sensitivity and 60.7% specificity (**Figure 3**). Logistic regression analysis showed that participants with choline ≥331.7 mg/d had 1.29-fold increased odds for DM after adjustment for multiple covariates (**Table 3**).

**Figure 3.**
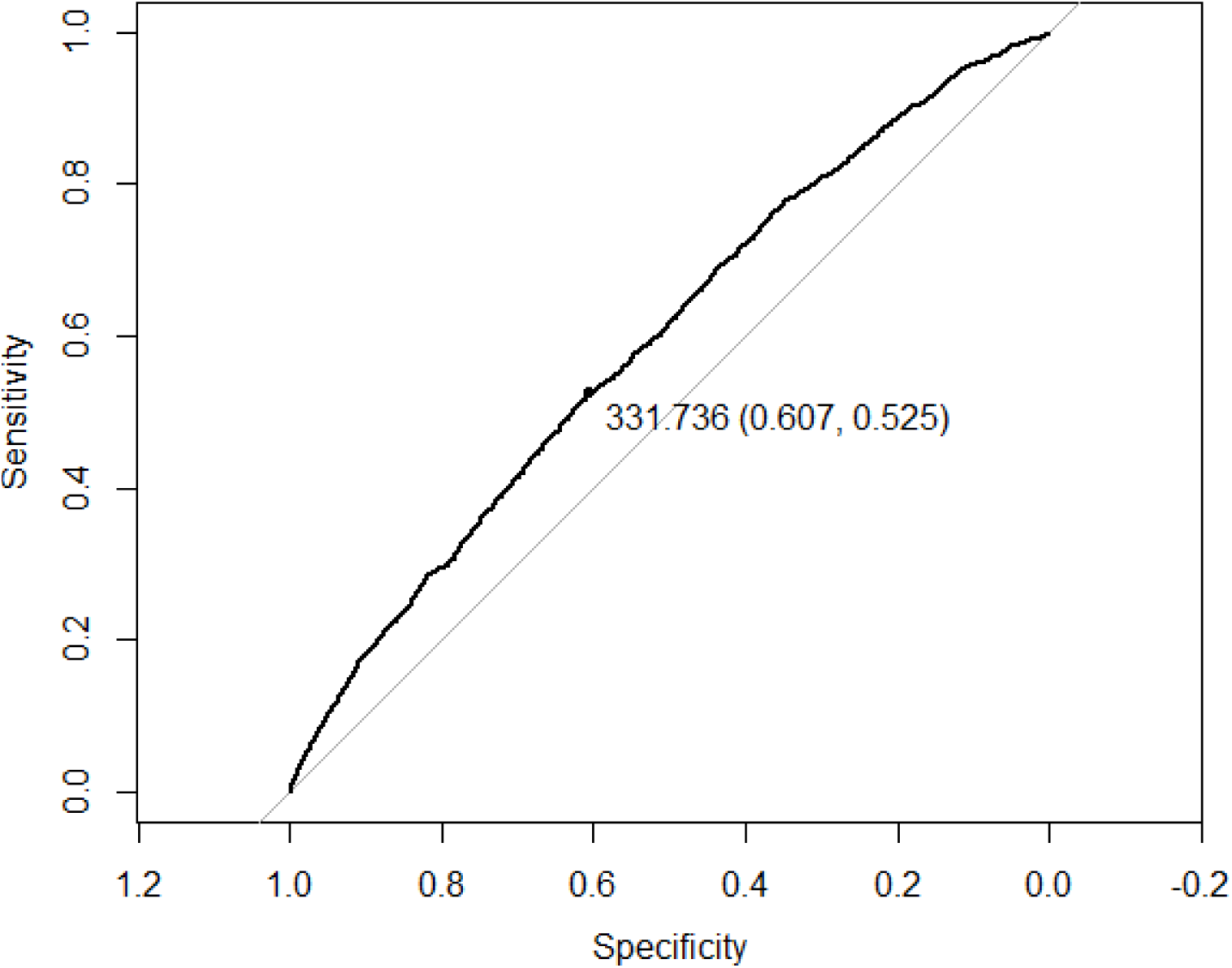
Receiver operating characteristic (ROC) curve and optimal cut-off of energy-adjusted choline intake for predicting diabetes

**Table 3.** Odds ratios and 95% confidence intervals for diabetes according to the optimal cut-off of energy-adjusted choline intake

| Dietary choline | Diabetes/n | Prevalence (%) | Age- and sex-adjusted model | Full-adjusted model<sup>2</sup> |
| --- | --- | --- | --- | --- |
| Optimal cut-off<sup>1</sup> |  |  |  |  |
| Below | 519/5083 | 10.2 | 1 (Ref) | 1 (Ref) |
| Equal or above | 574/3538 | 16.2 | 1.44 (1.26, 1.64) | 1.29 (1.11, 1.49) |

1 The optimal cut-off value of dietary choline is 331.7 mg/d.
2 Full-adjusted model adjusted for age, sex, ethnicity, education levels, family income levels, smoking status, drinking status, passive sedentary hours, body mass index, C-reactive protein, dietary energy, saturated fatty acids, polyunsaturated fatty acids, fiber, folate, vitamin B-6, and vitamin B-12 intakes.

## DISCUSSION

In this cross-sectional study on a nationally representative sample of US adults, we found that a higher level of dietary choline intake was positively associated with increased odds for DM, resulting in a linear dose-response relationship. This relationship was independent of multiple potential covariates including demographics, lifestyles, CRP, nutrients involved in the choline metabolism, and others.

Our hypothesis is that that higher dietary choline intake may be associated with an increased risk of DM. This is based on several studies that reported positive associations between plasma TMAO and DM in humans. A study involving 191 patients who underwent percutaneous coronary intervention has found that log-transformed TMAO levels was positively associated with DM status (multivariate adjusted β =0.058, p=0.009) (9). Results from a case-control study including 1346 newly diagnosed cases of type 2 diabetes (T2DM) and 1348 age- and sex-matched controls have shown that the multivariable-adjusted ORs (95% CIs) for T2DM were 1.00 (reference), 1.38 (1.08,1.77), 1.64 (1.28, 2.09), and 2.55 (1.99, 3.28) (p for trend <0.001) from the lowest to the highest quartiles of plasma TMAO (10). More evidence supporting positive associations between TMAO and glucose tolerance or insulin resistance was also found in animal experiments (15-17).

As a major dietary precursor for TMAO, choline in the development and prevention of DM is worth studying. However, there are limited studies exploring the relationship between dietary choline and DM, and furthermore, existing evidence remains conflicting. A large prospective study pooling results from the Nurses’ Health Study (NHS) I and II as well as the Health Professionals Follow-up Study (HPFS) found that compared with people in the lowest quintiles of dietary phosphatidylcholine (a derivative of choline) intakes, the multivariate-adjusted relative risk (RR) and 95% CI for T2DM for those in the highest quintiles was 1.36 (1.26, 1.48) in NHS I, 1.35 (1.22, 1.50) in NHS II, 1.28 (1.14, 1.44) in HPFS, and 1.34 (1.27, 1.42) in the pooled analysis (11). In contrast, a recent study including 2332 men from the Kuopio Ischaemic Heart Disease Risk Factor Study (KIHD) in eastern Finland has shown an inverse relation between dietary choline intake and the risk of T2DM. Specifically, those with the highest choline intake quartile had a 25% (95% CI: 2%, 43%) lower relative risk compared with those with the lowest choline intake (12). Our findings are consistent with the NHS and HPFS conducted in the US, but are in contrast with the KIHD conducted in Finland. One possible explanation is that the dietary pattern differs from country to country, which leads to the varied composition of gut microbiota in populations from different countries. Because not all strains of bacteria can convert choline to trimethylamine, there is a substantial inter-population variation in the circulating TMAO concentrations in response to choline intake (18).

Our present study showed a linear relation between dietary choline levels and the odds of DM. We used ROC analysis and found that choline ≥331.7 mg/d was the optimal cut-off point for predicting DM. Although the current dietary guidelines for Americans did not provide the recommended dietary allowance (RDA) of dietary choline due to insufficient data (19), the implication of our findings is that dietary choline should not exceed 331.7 mg per 2000 kcal caloric intake for US adults to achieve potentially lower risks of DM.

Our study has several limitations. First, given the cross-sectional nature of the NHANES study, it is not feasible to determine a causal relationship between dietary choline and DM. Second, the lack of plasma TMAO data makes it impossible to further analyze the mediation effect of TMAO on the association between dietary choline intake and DM. Third, due to the lack of antibody data, we cannot distinguish between type 1 and type 2 DM. However, the most common type is type 2 DM, which accounts for 90% to 95% of all cases of DM in the US, and type 1 DM constitutes 5% to 10% of DM (20,21).

In conclusion, this study supports a positive and linear relationship between dietary choline intake and odds of DM in the US adult population. The present study emphasizes the role of dietary choline in the development of DM, but further studies are warranted to replicate our findings in other populations and elucidate the potential mechanisms.

## Data Availability

All data used in this manuscript are de-identified and freely available to the public.

https://wwwn.cdc.gov/nchs/nhanes/default.aspx

## REFERENCES

1. Virani SS, Alonso A, Benjamin EJ, et al. Heart Disease and Stroke Statistics-2020 Update: A Report From the American Heart Association. Circulation 2020;141:e139-e596.

2. Sarwar N, Gao P, Seshasai SR, et al. Diabetes mellitus, fasting blood glucose concentration, and risk of vascular disease: a collaborative meta-analysis of 102 prospective studies. Lancet 2010;375:2215–22.

3. Saeedi P, Petersohn I, Salpea P, et al. Global and regional diabetes prevalence estimates for 2019 and projections for 2030 and 2045: Results from the International Diabetes Federation Diabetes Atlas, 9(th) edition. Diabetes Res Clin Pract 2019;157:107843.

4. Zhou L, Feng Y, Yang Y, et al. Diet behaviours and hypertension in US adults: the National Health and Nutrition Examination Survey 2013-2014. J Hypertens 2019;37:1230–1238.

5. Salas-Salvadó J, Martinez-González MÁ, Bulló M, Ros E. The role of diet in the prevention of type 2 diabetes. Nutr Metab Cardiovasc Dis 2011;21 Suppl 2:B32-48.

6. Wang Z, Klipfell E, Bennett BJ, et al. Gut flora metabolism of phosphatidylcholine promotes cardiovascular disease. Nature 2011;472:57–63.

7. Heianza Y, Ma W, Manson JE, Rexrode KM, Qi L. Gut Microbiota Metabolites and Risk of Major Adverse Cardiovascular Disease Events and Death: A Systematic Review and Meta-Analysis of Prospective Studies. J Am Heart Assoc 2017;6.

8. Tang W, Bäckhed F, Landmesser U, Hazen SL. Intestinal Microbiota in Cardiovascular Health and Disease: JACC State-of-the-Art Review. J Am Coll Cardiol 2019;73:2089–2105.

9. Dambrova M, Latkovskis G, Kuka J, et al. Diabetes is Associated with Higher Trimethylamine N-oxide Plasma Levels. Exp Clin Endocrinol Diabetes 2016;124:251–6.

10. Shan Z, Sun T, Huang H, et al. Association between microbiota-dependent metabolite trimethylamine-N-oxide and type 2 diabetes. Am J Clin Nutr 2017;106:888–894.

11. Li Y, Wang DD, Chiuve SE, et al. Dietary phosphatidylcholine intake and type 2 diabetes in men and women. Diabetes Care 2015;38:e13-4.

12. Virtanen JK, Tuomainen TP, Voutilainen S. Dietary intake of choline and phosphatidylcholine and risk of type 2 diabetes in men: The Kuopio Ischaemic Heart Disease Risk Factor Study. Eur J Nutr 2020. https://doi.org/10.1007/s00394-020-02223-2

13. Zipf G, Chiappa M, Porter KS, Ostchega Y, Lewis BG, Dostal J. National health and nutrition examination survey: plan and operations, 1999-2010. Vital Health Stat 1 2013:1-37.

14. American Diabetes Association. Standards of medical care in diabetes--2010. Diabetes Care 2010;33 Suppl 1:S11-61.

15. Dumas ME, Barton RH, Toye A, et al. Metabolic profiling reveals a contribution of gut microbiota to fatty liver phenotype in insulin-resistant mice. Proc Natl Acad Sci U S A 2006;103:12511–6.

16. Gao X, Liu X, Xu J, Xue C, Xue Y, Wang Y. Dietary trimethylamine N-oxide exacerbates impaired glucose tolerance in mice fed a high fat diet. J Biosci Bioeng 2014;118:476–81.

17. Li X, Chen Y, Liu J, et al. Serum metabolic variables associated with impaired glucose tolerance induced by high-fat-high-cholesterol diet in Macaca mulatta. Exp Biol Med (Maywood) 2012;237:1310–21.

18. Cho CE, Taesuwan S, Malysheva OV, et al. Trimethylamine-N-oxide (TMAO) response to animal source foods varies among healthy young men and is influenced by their gut microbiota composition: A randomized controlled trial. Mol Nutr Food Res 2017;61:1600324. https://doi.org/10.1002/mnfr.201600324

19. U.S. Department of Health and Human Services and U.S. Department of Agriculture. 2015–2020 Dietary Guidelines for Americans. 8th Edition. December 2015. Available at http://health.gov/dietaryguidelines/2015/guidelines/.

20. Centers for Disease Control and Prevention. National Diabetes Statistics Report, 2017: Estimates of Diabetes and Its Burden in the United States. Atlanta, GA: Centers for Disease Control and Prevention, US Department of Health and Human Services; 2017.

21. Menke A, Orchard TJ, Imperatore G, Bullard KM, Mayer-Davis E, Cowie CC. The prevalence of type 1 diabetes in the United States. Epidemiology 2013;24:773–4.

